# Characterising viral clearance kinetics in acute influenza

**DOI:** 10.1101/2025.03.07.25323547

**Authors:** Phrutsamon Wongnak, Tim Seers, Podjanee Jittamala, Mallika Imwong, William HK Schilling, James A Watson, Nicholas J White

## Abstract

Pharmacometric assessment of antiviral efficacy in acute influenza informs treatment decisions and pandemic preparedness. We characterised natural viral clearance in acute influenza to guide phase II trial design using simulations based upon observed data. Standardized duplicate oropharyngeal swabs were collected daily over 14 days from 80 untreated low-risk Thai adults, with viral densities measured using qPCR. We evaluated three models to describe viral clearance: exponential, bi-exponential, and growth-and-decay. The growth-and-decay model provided the best fit, but the exponential decay model was the most parsimonious. The median viral clearance half-life was 10.3 hours (interquartile range [IQR]: 6.8–15.4), varying by influenza type: 9.6 hours (IQR: 6.2–13.0) for influenza A and 14.0 (IQR: 10.3–19.3) hours for influenza B. Simulated trials using parameters from the exponential decay model, showed that 148 patients per arm provide over 90% power to detect treatments accelerating viral clearance by 40%. Variation in clearance rates strongly impacted the power; doubling this variation would require 232 patients per arm for an antiviral with a 60% effect size. A sampling strategy with four swabs per day reduces the required sample size to 81 per arm while maintaining over 80% power. We recommend this approach to assess and compare current anti-influenza drugs.

## Background

Despite extensive annual vaccination and widely available antiviral treatments, influenza virus continues to cause significant global morbidity and mortality. The concerns over the health threats posed by seasonal influenza are heightened by the threat of zoonotic pandemic influenza such as the Highly Pathogenic Avian Influenza (HPAI A/H5N1). Typically, viral loads in the upper respiratory tract in acute influenza peak around the second day of symptomatic infection, before declining over the following 3-5 days [1–3]. Antivirals effective against influenza, which include the neuraminidase inhibitors (zanamivir, oseltamivir and peramivir), nucleoside analogues (favipiravir) and cap-dependent endonuclease inhibitors (baloxavir) are given with the aim of reducing viral replication and attenuating illness, thereby reducing the risk of both progression to severe disease and onward virus transmission.

For the main antivirals currently in use (i.e. oseltamivir, peramivir, zanamivir, baloxavir) there is low certainty of evidence that they reduce severe outcomes such as hospitalisation and death [4]. This is because most seasonal influenza resolves uneventfully without treatment, and so very large studies in high risk sub-groups (e.g. the elderly or immunocompromised) are needed to evaluate or compare drugs in terms of prevention of hospitalisation and death. An alternative approach is to compare relative antiviral efficacies using measures of in-vivo viral clearance. Although influenza virus clearance has been measured in several drug assessments, in most studies the relatively inaccurate variable of *time to viral clearance* has been the primary outcome. This is a poor pharmacodynamic measure as it depends on the initial viral density, the frequency of measurement and the accuracy of detection at low densities, in addition to the magnitude of the antiviral effect [5].

Measuring the *rate of viral clearance* is a better approach and has been successfully used to evaluate and compare the antiviral efficacy of treatments for acute COVID-19 [6]. Using a simulation approach based on limited serial viral density measurements from naturally infected COVID-19 patients, Watson et al. (2022) [5] demonstrated that comparing viral clearance rates requires substantially fewer participants for a reliable comparison with a high statistical power than the trials using clinical endpoints, making this method well suited for phase II studies.

In this study, we characterise the viral clearance kinetics in untreated patients with early influenza infection from an ongoing adaptive platform trial (AD ASTRA, NCT05648448) and then use a simulation approach to determine the optimal sampling strategy and sample size needed to achieve sufficient statistical power for phase II pharmacometric clinical trials for influenza.

## Methods

### Participants

This study used data from an ongoing, open-label, multicentre phase II pharmacometric platform trial of antivirals for early influenza infection (AD ASTRA, NCT05648448), which is actively recruiting participants in Thailand, Laos, Brazil and Nepal. Following fully informed written consent, previously healthy patients aged 18-60 years with early symptomatic influenza (≤ 4 days) and a positive rapid diagnostic test or qPCR for influenza A or B virus (cycle threshold (CT) ≤ 30) were randomised initially to one of three antiviral treatment arms (favipiravir, oseltamivir, or baloxavir) or to a no study drug arm. Details on the inclusion and exclusion criteria, as well as clinical and laboratory procedures, are provided in the supplementary appendix (pp 2). Analyses were conducted in a modified intention-to-treat population (mITT), defined as patients whose average baseline viral density at enrollment (Day 0) exceeded 250 genomes/mL.

The goal of this analysis is to describe the influenza virus clearance kinetics in patients who had not received antiviral treatments. Therefore we included only data from patients in the no study drug arm. Due to the availability of qPCR data, only data from participants recruited in Thailand are reported here.

The trial was approved by the Oxford University Tropical Research Ethics Committee (Oxford, UK: OxTREC 6-23) and The Faculty of Tropical Medicine Ethics Committee, Mahidol University (Bangkok, Thailand: FTMEC 22-082).

### qPCR

The density of influenza virus genomes in the throat was determined from a validated qPCR assessment of daily serial oropharyngeal swab eluates taken on days 0–7 and 14 post-randomisation (4 samples on day 0 and 2 samples per day at subsequent time points). The TaqPath™ one-step RT-qPCR assay and custom complex assay 1TFS-2QAP-CCU002NR (Applied Biosystems, Thermo Fisher Scientific, Waltham, MA, USA) quantitated viral loads (RNA copies per mL). This multiplexed real-time PCR method detects influenza A, B, and human RNase P genes in a single reaction. To quantify viral loads, the ATCC^®^ VR-95DQ A/Puerto Rico/8/1934 (H1N1) strain for influenza A and the ATCC^®^ VR-1804DQ™ B/Florida/4/2006 strain for influenza B were used.

### Models of viral clearance kinetics

Individual patient viral density data were initially fitted to three phenomenological models describing viral clearance kinetics: exponential decay (log-linear), a bi-exponential decay, and an exponential growth-and-decay. Individual model parameters are assumed to follow a log-normal distribution.

The exponential decay model assumes that the viral density decreases exponentially at a single constant rate:

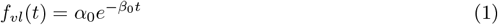

where *f*_*vl*_(*t*) represents the viral density (in genomes/mL) at time *t* days post-randomisation, and *α*_0_ > 0 denotes the baseline viral density (intercept) and *β*_0_ > 0 denotes the viral clearance rate.

The bi-exponential decay model assumes that the viral density decreases exponentially at two constant rates, dividing the kinetics into an initial rapid clearance phase and a later slow clearance phase:

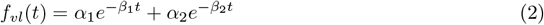

where *α*_1_ > *α*_2_ > 0 and *β*_1_ > *β*_2_ > 0 represent the baseline viral densities of each component and the viral clearance rates, respectively. The ratio *α*_2_*/α*_1_ determines the viral density at which the clearance transitions into the later slower terminal clearance phase.

The exponential growth-and-decay model [5, 7] divides the viral kinetics into two phases: an initial growth phase followed by a decay phase:

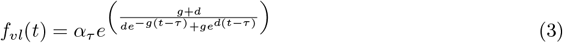

where *α*_*τ*_ represents the peak viral density occurring at time *t* = *τ* . The parameters *g* and *d* represent the growth and decay rates (in log_10_ genomes/mL per day), respectively. This accomodates patients whose infection is still expanding when they present to medical attention.

### Model fitting

We fit all models to data under a hierarchical Bayesian framework. Previous studies have demonstrated non-Gaussian variation in viral load estimates from oropharyngeal and nasopharyngeal swabs [5, 7, 8]. We modelled the observation error on the logarithmic scale using a *t*-distribution with unknown degrees of freedom *ν*:

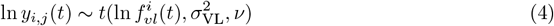

where *y*_*i,j*_(*t*) represents the *j*^*th*^ replicated qPCR measurement for viral density in oropharyngeal eluates of individual *i* at time *t* (in genomes/mL), while 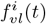 (*t*) is the model predicted viral density of individual *i* at time *t*. 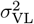 represents the variance of the observation error (intra-individual variation), this includes measurement error resulting in variation in swabbing technique, viral transport medium, and qPCR measurement.

Inter-individual variation in viral clearance kinetics are characterised by multiplicative changes in the model parameters, using as a *k*-dimensional vector 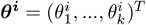, where *k* is the number of model parameters assumed to vary across patients. For example, inter-individual variation for the exponential decay model is written as:

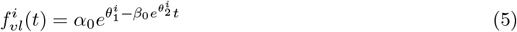

where *k* = 2, and 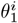 and 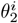 represent the random effects of individual *i* on the intercept (*α*_0_) and the slope (*β*_0_), respectively. In addition, under the exponential decay model, the viral clearance half-life, 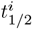, for each patient *i* can be calculated as 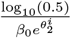.

Similarly to the exponential decay model, we parameterised individual random effects on the bi-exponential decay and exponential growth-and-decay models as shown in Eq. 6 and Eq. 7, respectively.

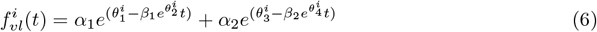

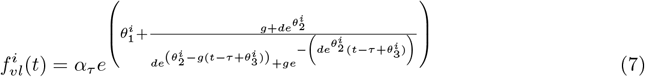

To aid convergence during parameter estimation, the random effect on the growth rate parameter *β*_1_ in the exponential growth-and-decay model was omitted from Eq. 7.

### Prior distributions

Weakly informative prior distributions were assigned for all parameters, with detailed descriptions provided in the appendix (pp. 4). Choices of prior distributions were informed by previously published SARS-CoV-2 clearance kinetics parameters [6, 9–13]. A multivariate normal prior distribution was specified for the individual random errors ***θ***^***i***^:

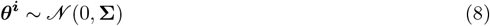

where **Σ** is the variance-covariance matrix governing inter-individual variation across the model parameters.

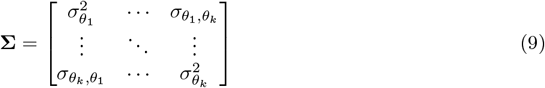

For parameter estimation, the variance-covariance matrix **Σ** was parameterised through its Cholesky LKJ decomposition, with a uniform prior distribution assigned to the correlation matrix *L*, and the variance terms expressed as standard deviations 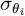.

### Parameter estimation and model comparison

Since we are aiming to characterise antiviral efficacy in reducing viral densities during the early stages of infection, viral clearance kinetics were characterised using serial viral density data collected up to 5 days post-randomisation as informed by a previous study of similar design in SARS-CoV-2 [14]. Viral clearance kinetics were also characterised using data from 7 days post-randomisation as a sensitivity analysis. Viral densities below the lower limit of detection (*LOD*), corresponding to a CT value of 40, were treated as left censored. As a result, the likelihood of the data *x* given the model parameters *θ* is 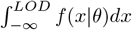, where *f* (*·*| *θ*) is the model likelihood. The analyses included three data sets: the complete data set and two sub-data sets for influenza A and influenza B.

All statistical analyses in this study were performed in R version 4.4.1. The hierarchical Bayesian models were implemented in *stan* and fitted using Hamiltonian Monte Carlo with the *rstan* package version 2.32.6 [15]. Model comparison was performed using leave-one-out cross-validation, as implemented in the *loo* package version 2.8.0 [16], along with residuals analysis and posterior predictive checks.

### Optimal trial design

#### Modelling antiviral efficacy

The AD ASTRA study is a Phase II clinical trial that uses virological endpoints to evaluate the antiviral activity of treatments in previously healthy patients with acute influenza. We define antiviral efficacy as the proportional change in the rate of influenza virus clearance in the patients allocated to an intervention arm relative to those in the no study drug arm, parameterised from the exponential decay model, the most parsimonious model. This is given as:

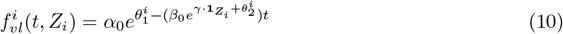

where *Z*_*i*_ is the randomised treatment allocation for individual *i* (1 for treatment arm, 0 for no study drug arm) and 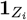 is the indicator function that takes the value 1 if *Z*_*i*_ = 1 and 0 otherwise. The parameter *γ* represents the effect size of the antiviral treatment. For example, an antiviral treatment *Z*_*i*_ = 1 results in a 20% increase in the viral clearance rate relative to the no study drug arm when *e*^*γ*^ = 1.2, i.e. *γ* = ln(1.2). The treatment has no antiviral efficacy when *e*^*γ*^ = 1.0 (i.e. *γ* = 0).

### Simulating clinical trials for influenza

To evaluate the impact of the number of patients per arm on the statistical power to detect a given effect size, a series of hypothetical trials were simulated. Daily serial viral density data were simulated using the posterior predictive distribution of the exponential decay model fitted to the influenza A data. We focused on influenza A kinetics to prevent variation in influenza type proportions across sites and periods from affecting the power estimates. Trials were simulated in which oropharyngeal eluate samples were collected twice daily over 5 days post-randomisation, providing a total of 12 viral density measurements per patient. The impact of recruiting varying numbers of participants per arm (10, 20, 40, 80, 160, 320, and 640 patients) was assessed when comparing hypothetical antiviral treatments with effect sizes (acceleration in clearance relative to no drug) of 0%, 20%, 40%, 60%, 80%, and 100%, corresponding to *γ* = ln(1.0), ln(1.2), ln(1.4), ln(1.6), ln(1.8), ln(2.0), respectively.

One hundred simulations were performed for each combination of effect size and number of patients per arm. For effect sizes of 20%, 40%, 60%, 80%, and 100%, statistical power was defined as the proportion of simulations (out of 100) in which the probability of the estimated antiviral efficacy 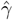 from its posterior distribution being less than or equal to 0 was less than 0.025 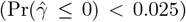, indicating the correct rejection of the null hypothesis at a level of 2.5%. For an effect size of 0%, the same probability, 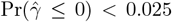, represents the type I error. Additionally, the expected sample size required to achieve 80% power was estimated using logistic regression.

In addition, estimation error, defined as the difference between the median posterior estimate of 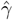 and the true parameter *γ*, expressed as 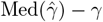, was calculated to assess the accuracy of the effect estimate. High accuracy implies an estimation error close to 0, with low dispersion in the estimation error across all simulations, respectively.

#### Sensitivity analysis and alternative sampling strategies

As a sensitivity analysis, we conducted additional simulated trials to evaluate the impact of three sources of variation in the serial viral density data on statistical power: 1) inter-individual variation in the intercept 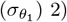 inter-individual variation in the slope 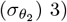 observational error (*σ*_VL_). In addition to the same combinations of patient numbers per arm and antiviral efficacies, daily serial viral density data were simulated with the values of the three variations set to 0.5x, 1.0x, and 2.0x, each evaluated individually to estimate the effect size of antiviral efficacies.

We also examined how alternative sampling strategies affected statistical power and estimation errors, varying: 1) the sampling schedule — every day (6 days), every other day (3 days), or only on the first and last day (2 days); and 2) the number of swabs per day — either 1, 2, or 4 swab(s).

In addition, we evaluated the impact of the simulated data-generating process by conducting simulations using the posterior predictive distributions of the bi-exponential and growth-and-decay models to estimate power and type I error. The antiviral treatment (*γ*) was parameterised to modify the initial clearance rate (*β*_1_) in the bi-exponential model (Equation 11) and the decay rate (*d*) in the growth-and-decay model (Equation 12), respectively.

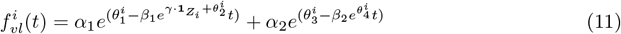

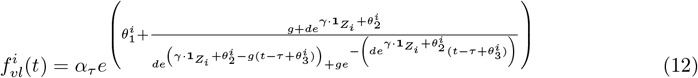

Statistical power and estimation errors were calculated over 50 simulations for these ad hoc analyses.

## Results

### Participants

Of the 355 patients assessed for eligibility for the AD ASTRA trial at the Thailand site between 22 February 2023 and 31 December 2024, 91 were randomised to the no study drug arm, of whom 70 had influenza type A and 21 influenza type B (Figure 1). Of these, 11 patients had average oropharyngeal eluate viral densities at enrollment (Day 0) below 250 log_10_ genomes/mL, leaving a total of 80 patients in the modified intention-to-treat (mITT) population (60 influenza A, 20 influenza B). Of these, 4 patients with influenza A infection did not complete the trial due to consent withdrawal within two days post-randomisation. None of the participants reported serious adverse events and all recovered uneventfully.

**Figure 1:**
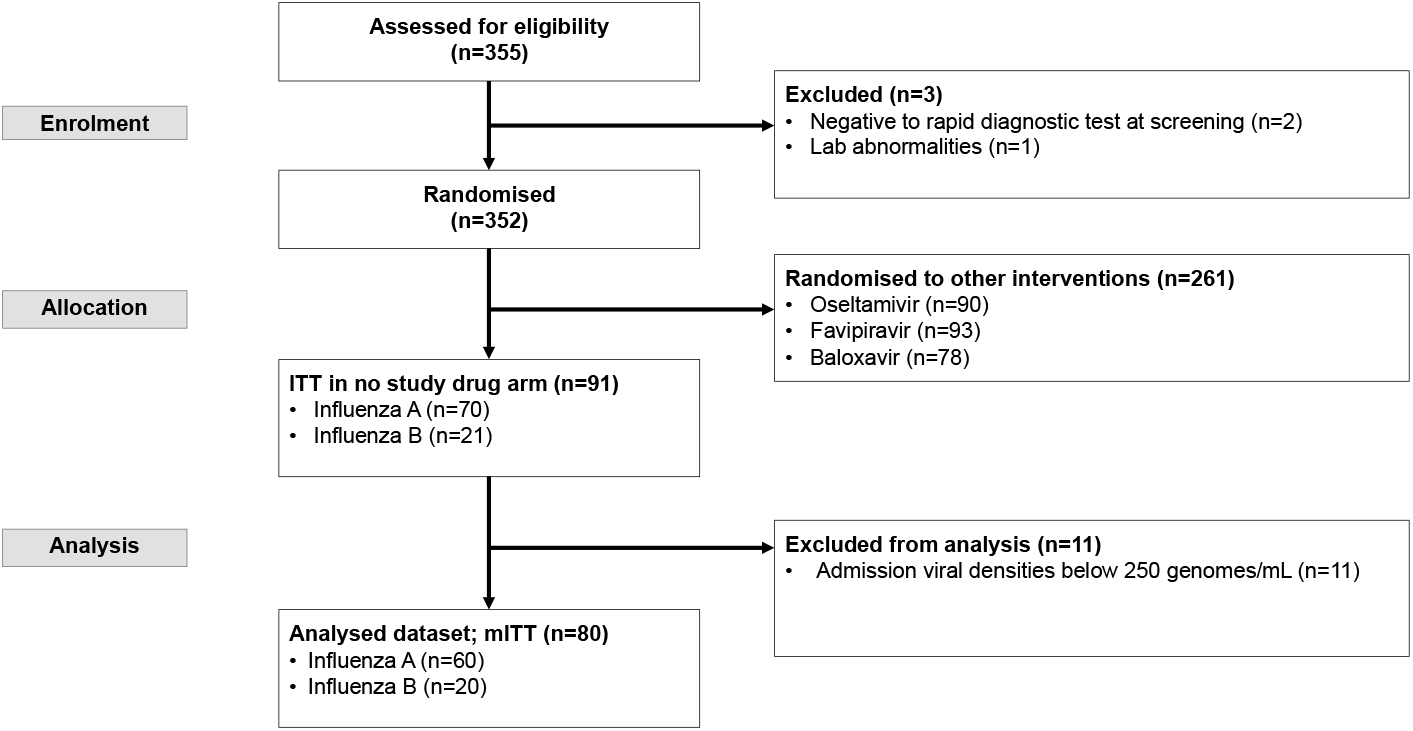
Study CONSORT diagram for characterizing influenza clearance kinetics in patients randomised to no study drug arm from AD ASTRA trial between 22 February 2023 to 31 December 2024.

Among the 80 mITT patients, 41 (58%) were female, with a median age of 31 years (interquartile range: 26 to 40) (Table 1). Of these, 33 (41%) were enrolled in the study two days after the onset of their symptoms. Only 6% (5/80) of the patients had been vaccinated.

**Table 1:** Baseline patient characteristics in modified intention-to-treat populations.

| Characteristics | Influenza A<br>(n = 60) | Influenza B<br>(n = 20) | Total<br>(n = 80) |
| --- | --- | --- | --- |
| Female | 33 (58%) | 8 (57%) | 41 (58%) |
| Age<sup>a</sup> (year) | 32 (25 to 40) | 32 (28 to 40) | 31 (26 to 40) |
| Weight<sup>b</sup> (kg) | 67 (13) | 66 (12) | 67 (13) |
| Body mass index<sup>b</sup> | 24 (4) | 25 (5) | 25 (4) |
| Admission oropharyngeal eluate viral density<sup>b</sup> (log<sub>10</sub> genomes/mL) | 4.7 (1.3) | 5.5 (1.4) | 4.9 (1.3) |
| Day(s) post symptom onset<sup>a</sup> | 2 (1 to 3) | 2 (2 to 3) | 2 (1 to 3) |
| Vaccination history: |  |  |  |
| previously received influenza vaccine | 4 (7%) | 1 (5%) | 5 (6%) |
| Not vaccinated | 56 (93%) | 19 (95%) | 75 (94%) |

Unless specified, data are presented as number with percentage; <sup>a</sup>median with interquartile range; <sup>b</sup>mean with standard deviation.

### Viral clearance kinetics

After excluding 11 patients with admission oropharyngeal eluate viral densities below 250 genomes/mL, the average admission viral density was 4.9 (SD = 1.3) log_10_ genomes/mL (Table 1). Patients with influenza type A infections had lower admission viral densities (4.7 log_10_ genomes/mL, SD = 1.3) compared to those with influenza type B (5.5 log_10_ genomes/mL, SD = 1.4) (Table 1, Figure 2). For influenza A infections, >50% of patients had undetectable viral densities by day 4 (i.e. CT values of 40 in all swab samples); for influenza B >50% of patients had undetectable viral densities by day 6 (Figure 2; Table S1). By Day 14, detectable viral densities were still observed in 2% of patients with influenza A and 15% for influenza B.

**Figure 2:**
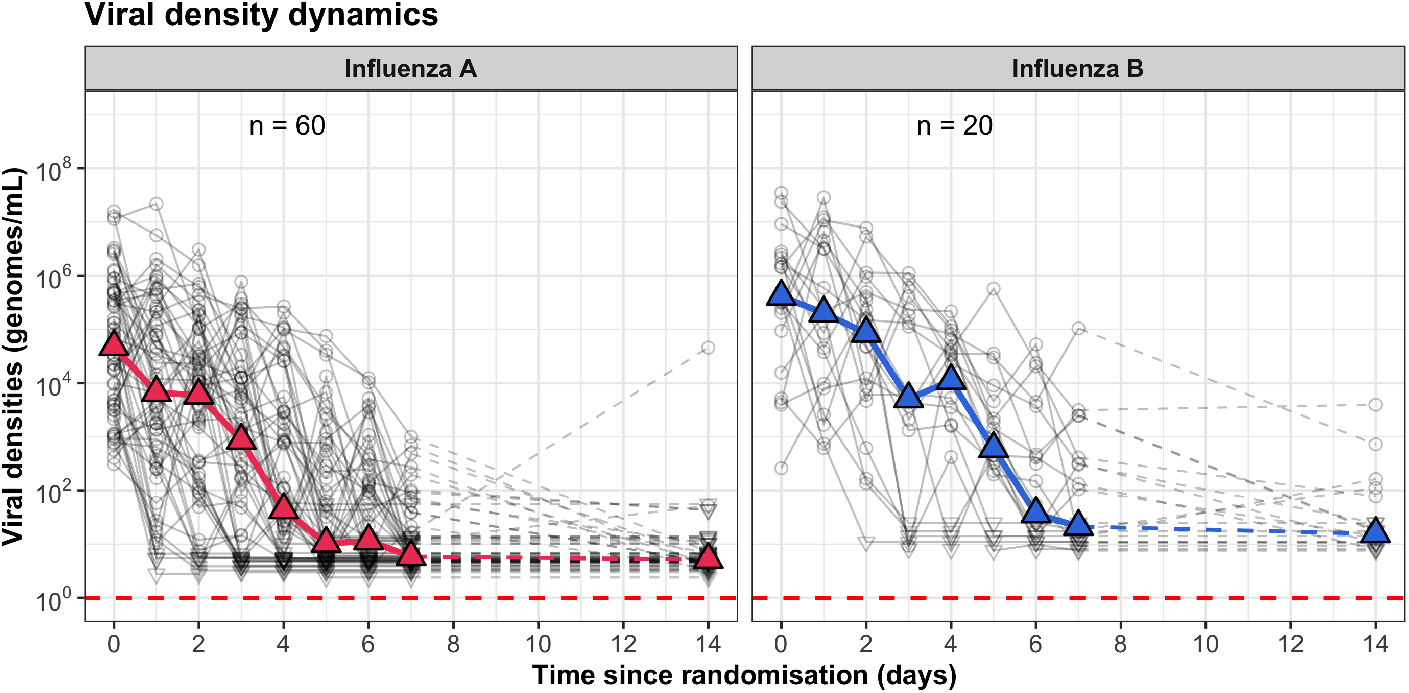
Serial influenza eluate viral densities (genomes/mL) measured from oropharyngeal swabs. Grey points and lines represent the daily average viral densities for individual patients. Values below the limit of quantification are shown as inverted triangles. Filled triangles and coloured lines indicate the median values.

Three phenomenological models - exponential decay, bi-exponential decay, and growth-and-decay — were fitted to the serial viral density data over 5 days post-randomisation (total of 14 swabs per patient). All three models demonstrated a good fit, with residuals centred around 0, with greater deviations from zero were observed at later time points, primarily because of left-censored data points (Figure 3). Details of serial viral density data and the corresponding model fits for individual patients are provided in the supplementary materials (pp 5).

**Figure 3:**
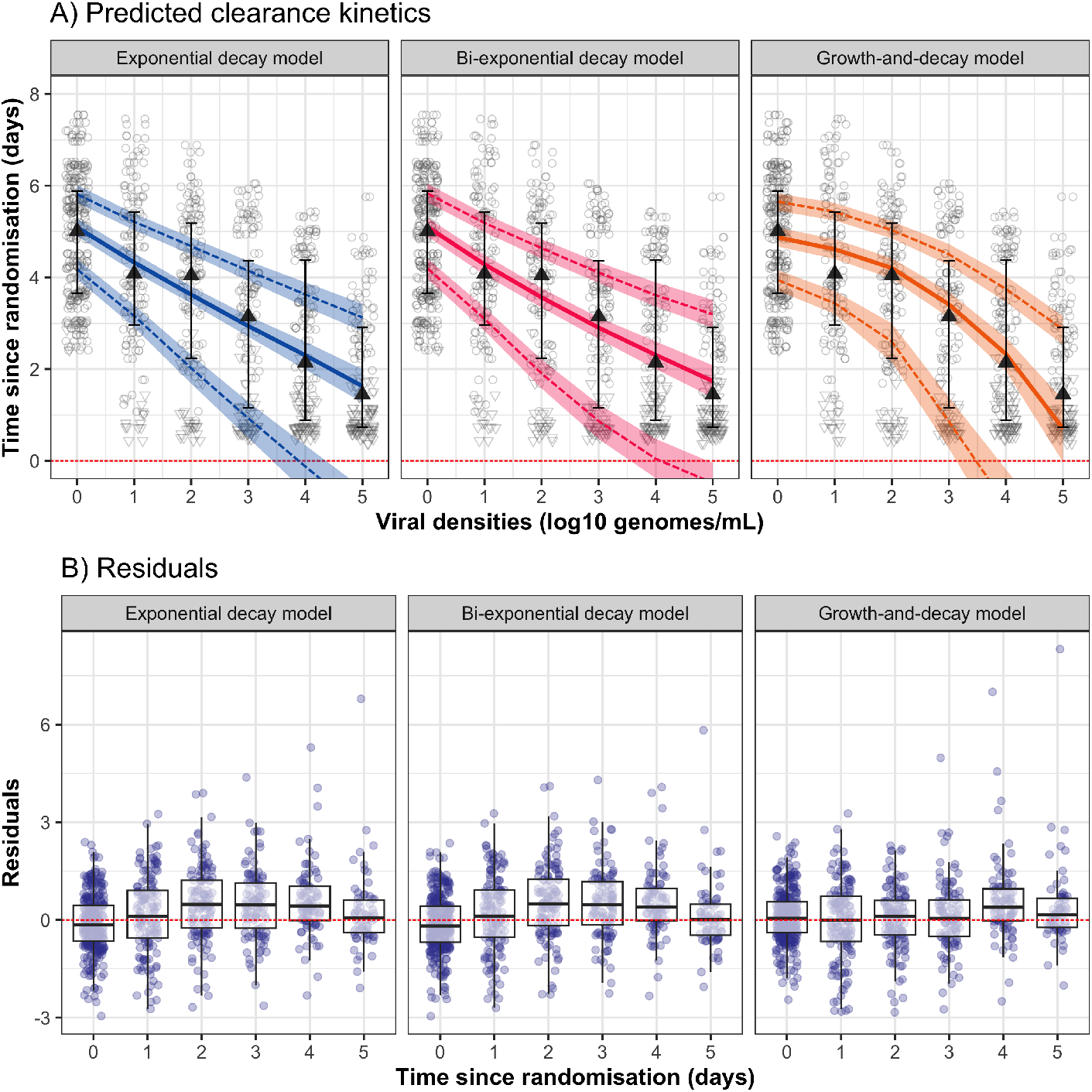
Oropharyngeal influenza viral clearance kinetics fitted to three phenomenological models (exponential decay, bi-exponential decay, and growth-and-decay models): A) Posterior predictions of population median (coloured solid line) and first and third quartiles (coloured dashed lines) compared with observed daily serial viral densities (triangles with whiskers). Shaded areas represent 95% credible intervals. Blank points represent the daily average viral densities of individual patients, with inverted triangles indicating left-censored values with a CT value of 40. The filled triangles and black solid line represent the median daily viral density of the recruited patients; B) Model residuals (in log10 genomes/mL) as a function of time since randomisation, calculated from model predictions for each individual patient.

The Pareto *k* diagnostic values indicate that more than 97% of the observations fall within the reliable range of the three models (*k* ≤ 0.67), suggesting robust model performance. The expected log-predictive density (ELPD) was lowest for the growth-and-decay model (ELPD = -1553; 95% CI: -1616 to -1489), identifying it as the best-fitting model. However, there were only small differences in ELPD relative to the bi-exponential (-51, 95%CI: -79 to -23) and the exponential decay models (-47; 95% CI: -71 to -22).

The growth-and-decay model predicted that 64% (51 out of 80) of the recruited patients had reached their peak viral densities before randomisation, suggesting they were already in the decay phase of viral kinetics upon admission. Only 23% (18 out of 80) reached peak densities after Day 1 post-randomisation. Additionally, predictions from the bi-exponential model showed that in 95% (76/80) of the patients, the viral densities entering the slow clearance phase, indicated by the parameter *α*_2_ from Eq. 6, were below 100 genomes/mL (near the limit of detection). This suggests that viral clearance during the 5 days after randomisation is primarily driven by the initial rapid clearance phase, approximated by an exponential decay model.

Based on the single exponential decay model, the average viral clearance rate for influenza over the first 5 days after randomisation was estimated at 0.7 log_10_ genomes/mL per day (95% credible interval [CrI]: 0.6 to 0.9; Figure 4A; Table S2). This corresponds to an average viral clearance half-life of 10.1 hours (95% CrI: 8.4 to 12.1). As a sensitivity analysis, when extending the analysis to 7 days post-randomisation, the average clearance rate was estimated as 0.8 log_10_ genomes/mL per day (95% CrI: 0.7 to 0.9), equivalent to the average viral clearance half-life of 9.1 hours (95% CrI: 8.6 to 9.5). Within the population, the median viral clearance half-life was 10.3 hours, with an interquartile range (IQR) of 6.8 to 15.4 hours (Figure 4B).

**Figure 4:**
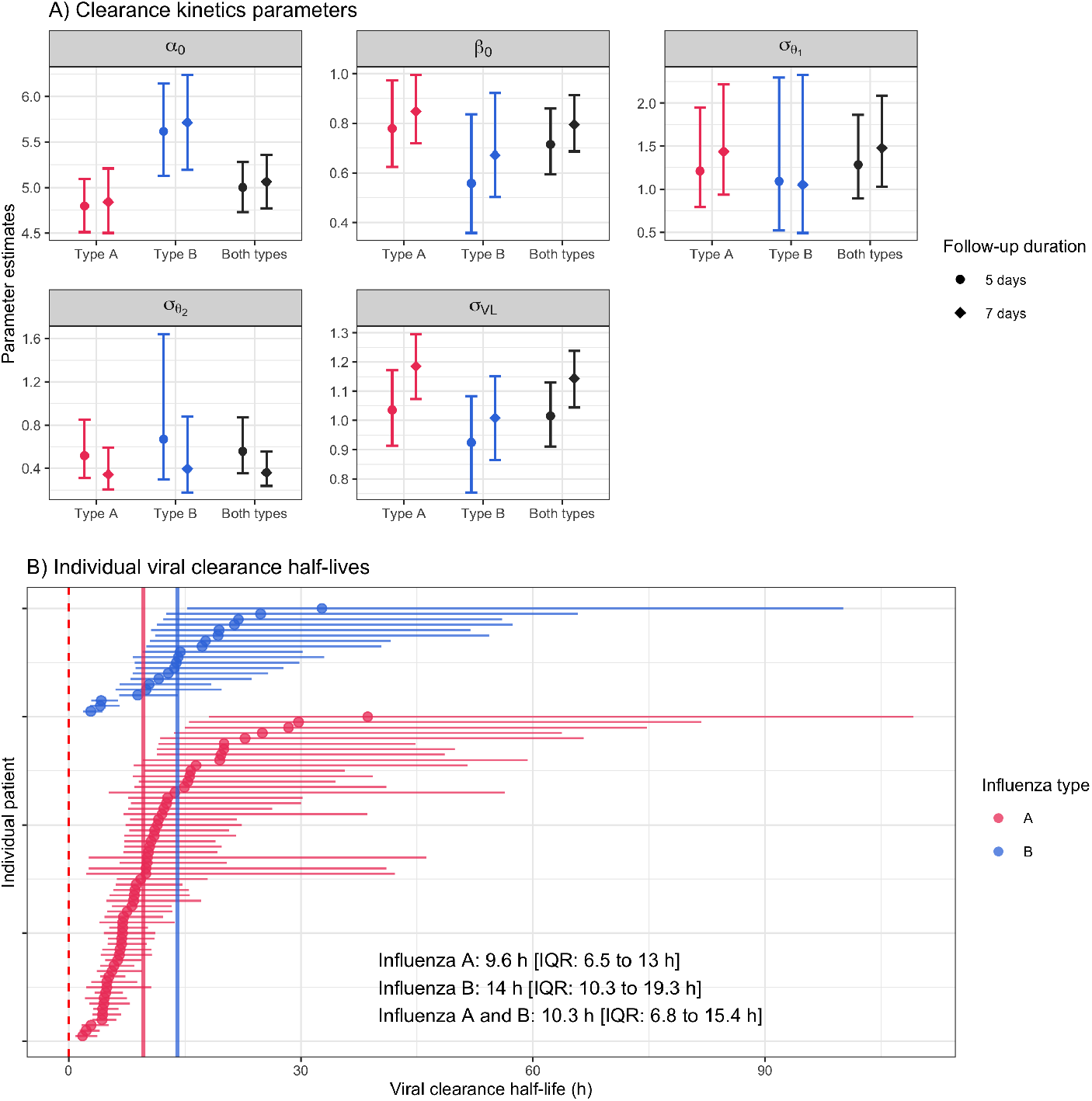
Panel A: Posterior distribution of clearance kinetics parameters for the exponential decay model. *α*_0_: population mean admission viral density (log_10_ scale); *β*_0_: population mean viral clearance rate; *σ*_VL_ observation error (log_10_ scale); 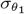 standard deviation of inter-individual variation in baseline viral densities *α*_*i*_ (log_10_ scale); 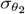 standard deviation of inter-individual variation on the clearance rate (multiplicative variation on ln scale). Parameters were estimated using the full dataset (black), influenza A (red), and influenza B (blue) subgroups. Points and error bars represent the median and 95% credible interval of the posterior distributions, respectively. Panel B: Individual viral clearance half-lives, with colours indicating influenza type (influenza A in red, influenza B in blue). Points and error bars represent the median and 95% credible intervals, respectively. Vertical solid lines indicate the median values for each influenza subgroup.

The observation error (*σ*_VL_), inter-individual variation in the intercept 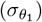, and inter-individual variation in the slopes 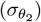 were estimated as 1.0 (95% CrI: 0.9 to 1.1), 1.3 (95% CrI: 0.9 to 1.9), and 0.6 (95% CrI: 0.4 to 0.9), respectively. The estimated covariance between the intercepts and the slopes 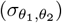 was -0.1 (95%CrI: -0.3 to 0.2). All variations are reported on the log_10_ scale.

Subgroup analysis showed clear differences in viral clearance kinetics between Influenza A and Influenza B 4A; Table S2). Influenza A was associated with lower admission viral densities (4.8 log_10_ genomes/mL; 95% CrI: 4.5 to 5.1) and a faster viral clearance (0.8 log_10_ genomes/mL per day; 95% CrI: 0.6 to 1.0) compared to Influenza B, admission viral densities (5.6 log_10_ genomes/mL; 95% CrI: 5.1 to 6.1) and clearance (0.6 log_10_ genomes/mL per day; 95% CrI: 0.4 to 0.8). The median viral clearance half-life for patients with Influenza A was 9.6 hours (IQR: 6.5 to 13.0), compared to 14.0 hours (IQR: 10.3 to 19.3) for those with Influenza B (Figure 4B). The observation error and inter-individual variation in both intercepts and slopes were similar for both influenza types and aligned with estimates from the full dataset (Figure 4A; Table S2).

The impact of using 7-day influenza clearance kinetics, by which time over 70% of swab samples were below the limit of detection, instead of 5-day post-randomisation data, was an inflation of observation error, a reduction in inter-individual variation of the slopes, and an overestimation of viral clearance rates.(Figure 4A; Table S2).

### Optimal design for influenza pharmacometric studies

Our simulation study, based on the estimated viral clearance kinetics for Influenza A, showed that recruiting 80 and 52 patients per arm, with analysable serial viral density data between days 0 and 5, in a treatment comparison would provide more than 90% and 80% power, respectively, to detect effect sizes of greater than 60% acceleration in viral clearance rate (Figure 5A, Tables S3 and S4). However, 148 patients per arm would be required to achieve 90% statistical power for the more moderate effect size of 40% acceleration. Type I error rate when testing a treatment with no antiviral effect (*γ* = 0), calculated from 100 simulations per condition, ranged from 0% to 0.06% (Figure 5A). Enrolling more than 80 patients per arm optimised further the accuracy and precision in estimating the effect size of antiviral activity. This is shown by the estimation errors centered around zero with reduced variation across all simulations (Figure S1).

**Figure 5:**
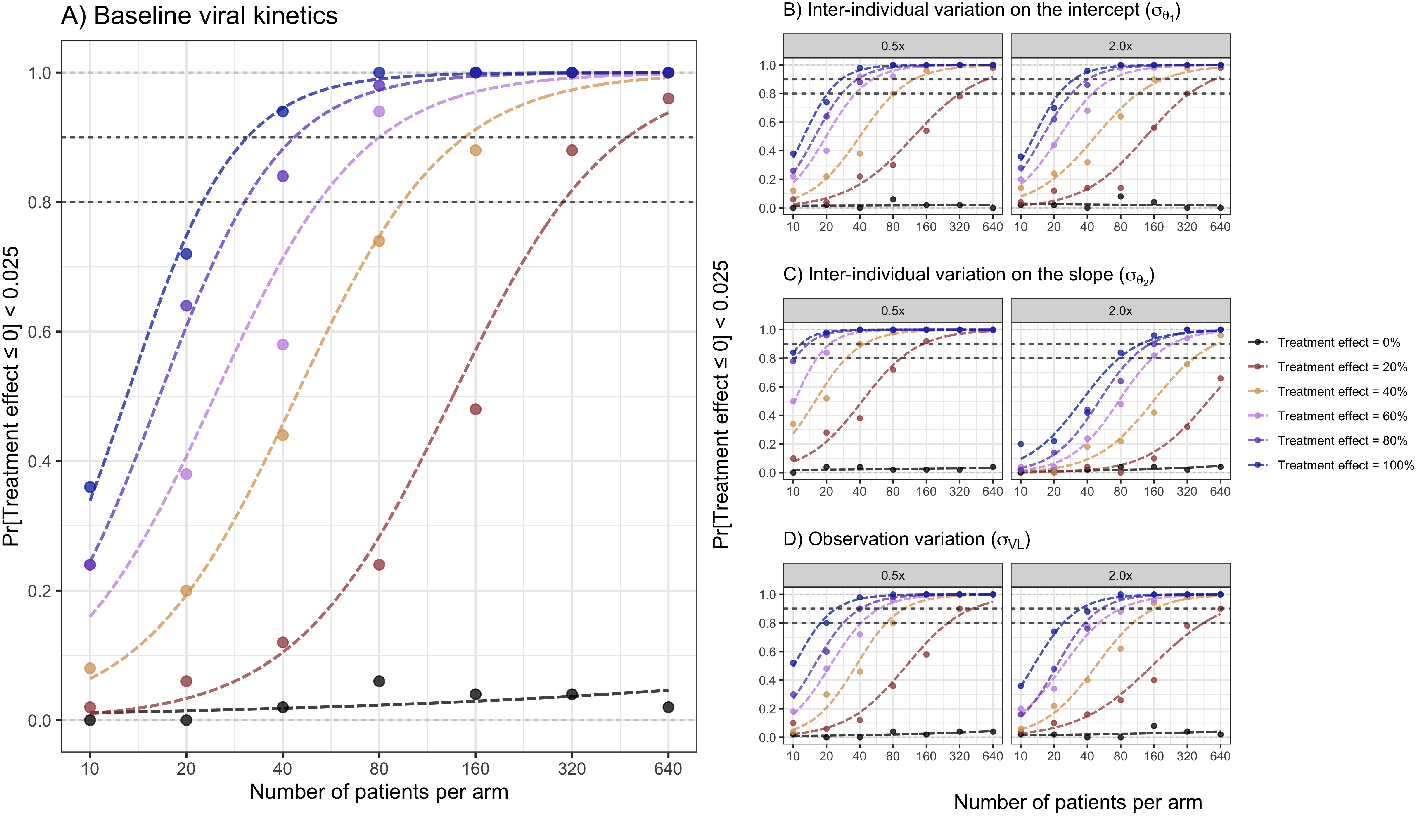
The relationship between the number of patients recruited per arm and statistical power (or type I error for an effect size of 0%), with colours indicating different treatment effect sizes. Logistic regression fits (dashed lines) are shown to estimate sample sizes required for 80% and 90% power. Panel A: Power was calculated across 100 simulations using viral clearance kinetics parameters from Influenza A infections (baseline). Panels B to D: Power was calculated across 50 simulations, adjusting for observation error (*σ*_VL_), inter-individual variation in the intercept 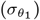, and inter-individual variation in the slope 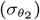, with adjustments of half (0.5x) and twice (2.0x) the parameter values of natural influenza A infections, one at a time.

The sensitivity analysis showed that statistical power, and the accuracy and precision of estimating the antiviral treatment effect size are influenced substantially by inter-individual variation in the clearance slope 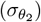. In contrast, they are less affected by observation error (*σ*_VL_) or inter-individual variation in the intercept 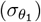 (Figures 5B to C, and S1).

In the simulation scenarios where the infection has twice the inter-individual variation in the slope 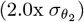, up to 232 patients per arm would be required to achieve more than 90% statistical power when testing an antiviral with ≥ 60% effect size (Figure 5C, Table S3). Recruiting fewer than 40 patients per arm would result in biased (underestimated) effect size estimates even when treatments had effect sizes as high as 100% acceleration in viral clearance (Figure S1C). In contrast, if the inter-individual variation in the slope is half that of natural influenza A infections 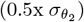, only 149 patients per arm would be needed to reach 90% power (Figure 5C, Table S3) with high accuracy and precision (Figure S1C) when testing an antiviral with a 20% effect size.

Alternative sampling strategies with two time points (day 0 and day 5) for viral density data slightly reduced statistical power, though the power to detect treatments with a 40% effect size remained above 80% when recruiting 113 patients per arm (Figure 6A, Table S4). Doubling the number of swab samples moderately improved statistical power, allowing the number of patients per arm to be reduced from 94 to 81 while maintaining power above 80% for detecting a 40% effect size (Figure 6B, Table S4).

**Figure 6:**
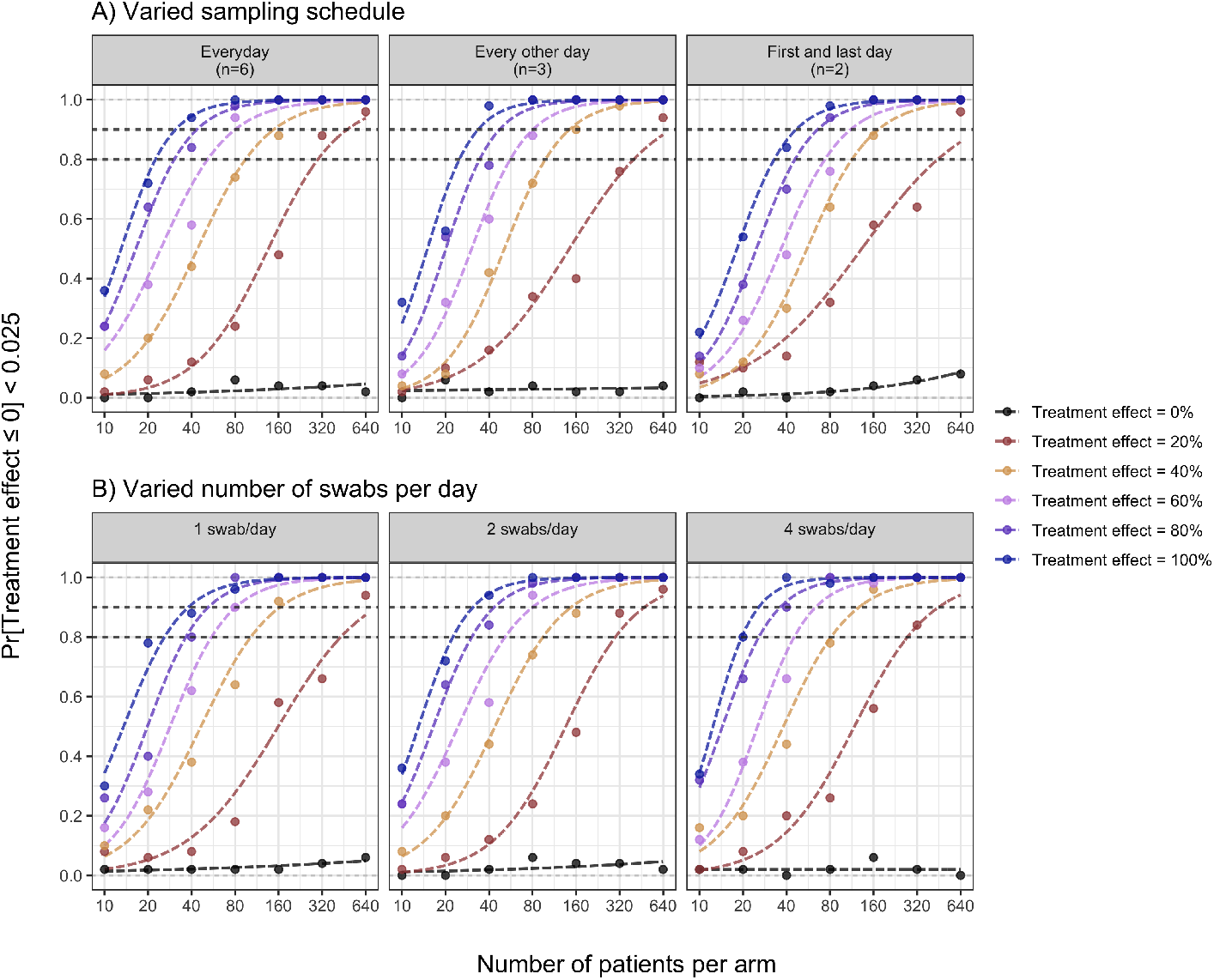
The relationship between the number of patients recruited per arm and statistical power (or type I error for an effect size of 0%) in testing the effectiveness of antiviral treatments in patients with acute influenza using alternative sampling strategies, with colours indicating different treatment effect sizes. Logistic regression fits (dashed lines) are shown to estimate the sample sizes required for 80% and 90% power. Panel A: Varying sampling schedules (every day, every other day, first and last day). Panel B: Varying number of swabs per day (1, 2, and 4 swabs).

Finally, when using a bi-exponential model as an alternative data-generating model, where antiviral treatments accelerate only the initial rapid decay phase, the estimated power was substantially affected under the current statistical framework (Figure S3). In contrast, using the growth-and-decay model, where treatments accelerate the decay phase, there was little impact on the power calculation.

## Discussion

Quantification of serial viral densities from oropharyngeal swab sample eluates is a simple and well-tolerated procedure which has allowed characterisation of viral clearance kinetics in acute SARS-CoV-2 infections. This simple sampling method facilitates high adherence rates in pharmacometric trials [6, 9–13], and provides important comparative antiviral efficacy estimates to inform policies and practices. Viral clearance data assessed from nasopharyngeal or oropharyngeal swab eluates is inherently “noisy”. Our simulations based on viral clearance rates measured in 80 naturally infected untreated adult individuals in Thailand indicate that inter-individual variation in the clearance slope primarily drives the sample size needed for characterising antiviral efficacy. The smaller the variation, the fewer participants that are required to achieve high statistical power.

Statistical power in the assessment of antiviral efficacy is defined as the probability of correctly rejecting the null hypothesis when the effect size is greater than a presepcified value. Achieving high power necessitates larger sample size per arm, especially when the true effect size of the antiviral treatment is small.

Our results indicate that statistical power, accuracy, and precision in testing antiviral effects are robust and more influenced by slope variation across the population than by observational error. Improving sampling techniques or schedules can moderately enhance statistical power by reducing observational error. Consistent with our power analysis on the SARS-CoV-2 trial [5], increasing the number of daily swabs improves statistical power and reduces the required number of patients per arm. However, the cost-benefit and feasibility of collecting more swabs should be carefully considered when designing the trial.

The optimum duration of follow-up in pharmacometric assessments is influenced by the background viral clearance rate [5, 14]. In COVID-19 infection the overall viral clearance profile is biphasic, but the initial phase is drug sensitive. Longer follow up increases “dilution” from the second phase and thereby reduces the discriminant power in comparing antivirals. In practical terms a follow-up duration of five days is superior to seven days. The same applies to influenza. Natural virus clearance rates are approximately similar for the two infections; SARS-CoV-2: half-life of 9.2 hours [14]; Influenza A: half-life of 9.6 hours estimated in this study; Influenza B: half-life of 14.0 hours. This suggests that a follow-up duration of 5 days or less would maximise statistical power for assessing anti-influenza treatments.

We observed a slower viral clearance rate and higher admission viral densities in Influenza B infections compared to Influenza A. This has been observed previously in other small studies of natural and volunteer challenge influenza infection [17, 18]. Data from ferret models suggest Influenza B virus preferentially antagonises the interferon driven innate immune response, and that this is associated with a delayed antibody-mediated response when compared to Influenza A infection [19]. Whilst this observation is consistent with previously published reports, we cannot be sure that the different limits of detection in our assay (and correspondingly different standard curves) has not contributed to the different clearance trajectories described. Additionally, inter-individual variation was approximately 40% higher in Influenza B, though this estimate is based on a relatively small sample size.

Interpretation of these results should take the following considerations into account. First, the clearance kinetics for influenza infections in this study are based on data from adults with mild disease in Thailand. These findings—particularly the differences between influenza A and B, and the estimated viral clearance rates and inter-individual variation—may not be generalisable to other settings or populations. When applying our findings to determine the required sample size for a treatment comparison, it is important first to characterise the baseline viral kinetics of the population and then adjust the sample size using the sensitivity analysis results. Second, the statistical framework used in this study is not designed to detect differences in responses to antiviral treatments across subgroups (e.g., influenza types, age, or sex) with high statistical power. The framework assumes that individuals within the same subgroup respond similarly to treatment. Under this framework, antiviral treatments are not modelled to influence inter-individual variation in the slope. Modifying the model parameterisation, such as including interaction terms between subgroups and treatment arms or adding a parameter to account for treatment effects on inter-individual variation, would help mitigate these limitations.

## Conclusions

This first detailed characterisation of virus clearance kinetics in natural acute influenza provides an evidence base for the design of pharmacometric drug evaluations. Daily duplicate sampling over five days with a sample size of 120 patients per arm provides high statistical power, accuracy, and precision to detect and estimate antiviral efficacy for treatments that accelerate viral clearance rates by ≥ 40%.

## Acknowledgements

We thank all the patients with influenza who volunteered to be part of the study. We thank the data safety and monitoring board (Tim Peto, André Siqueira, Sudha Basnet, and Panisadee Avirutnan), the trial steering committee (Nathalie Strub-Wourgaft, Martin Llewelyn, Deborah Waller, and Attavit Asavisanu), Sompob Saralamba and Tanaphum Wichaita (Mahidol Oxford Tropical Medicine Research Unit) for developing the Rshiny randomisation app. We also thank all the staff of the Clinical Trials Unit at Mahidol Oxford Tropical Medicine Research Unit for their excellent support with this project, and the Hospital for Tropical Diseases (Bangkok, Thailand), as well as those involved in sample processing at the Mahidol Oxford Tropical Medicine Research Unit and processing and analysis at molecular genetics laboratory and the malaria laboratory of the Faculty of Tropical Medicine, Mahidol University. We thank the Mahidol Oxford Tropical Medicine Research Unit Clinical Trials Support Group for data management, monitoring, and logistics, and the purchasing, administration, and support staff at Mahidol Oxford Tropical Medicine Research Unit.

## Authorship contributions

Conceptualization: PW, JAW, NJW. Supervision: WHKS, JAW, NJW; Methodology: PW, JAW. Formal analysis and visualization: PW. Data collection and investigation: PJ, MI, TS, WHKS. Writing—original draft: PW, TS. Writing—review and editing: all authors. Funding acquisition: WHKS, JAW, NJW.

## Data availability

All data and code necessary to reproduce the results in this analysis are openly available on GitHub https://github.com/psamonwong/Viral_clearance. All code and de-identified participant data required for replication of the study’s endpoints are openly accessible through Zenodo, as well as the study protocol and statistical analysis plan, from publication date onwards.

## Supplementary information

**Table S1:** Proportion of patients with viral densities below the limit of quantification in all swabs taken on each visit.

| Influenza type | Day 0 | Day 1 | Day 2 | Day 3 | Day 4 | Day 5 | Day 6 | Day 7 | Day 14 |
| --- | --- | --- | --- | --- | --- | --- | --- | --- | --- |
| Influenza type A | 1/60<br>(2%) | 9/57<br>(16%) | 12/56<br>(21%) | 20/56<br>(36%) | 31/56<br>(55%) | 40/56<br>(71%) | 39/55<br>(71%) | 49/56<br>(88%) | 55/56<br>(98%) |
| Influenza type B | 1/20<br>(5%) | 0/20<br>(0%) | 4/20<br>(20%) | 6/20<br>(30%) | 5/20<br>(25%) | 5/20<br>(25%) | 11/20<br>(55%) | 14/20<br>(70%) | 17/20<br>(85%) |
| All | 2/80<br>(2%) | 9/77<br>(12%) | 16/76<br>(21%) | 26/76<br>(34%) | 36/76<br>(47%) | 45/76<br>(59%) | 50/75<br>(67%) | 63/76<br>(83%) | 72/76<br>(95%) |

**Table S2:**
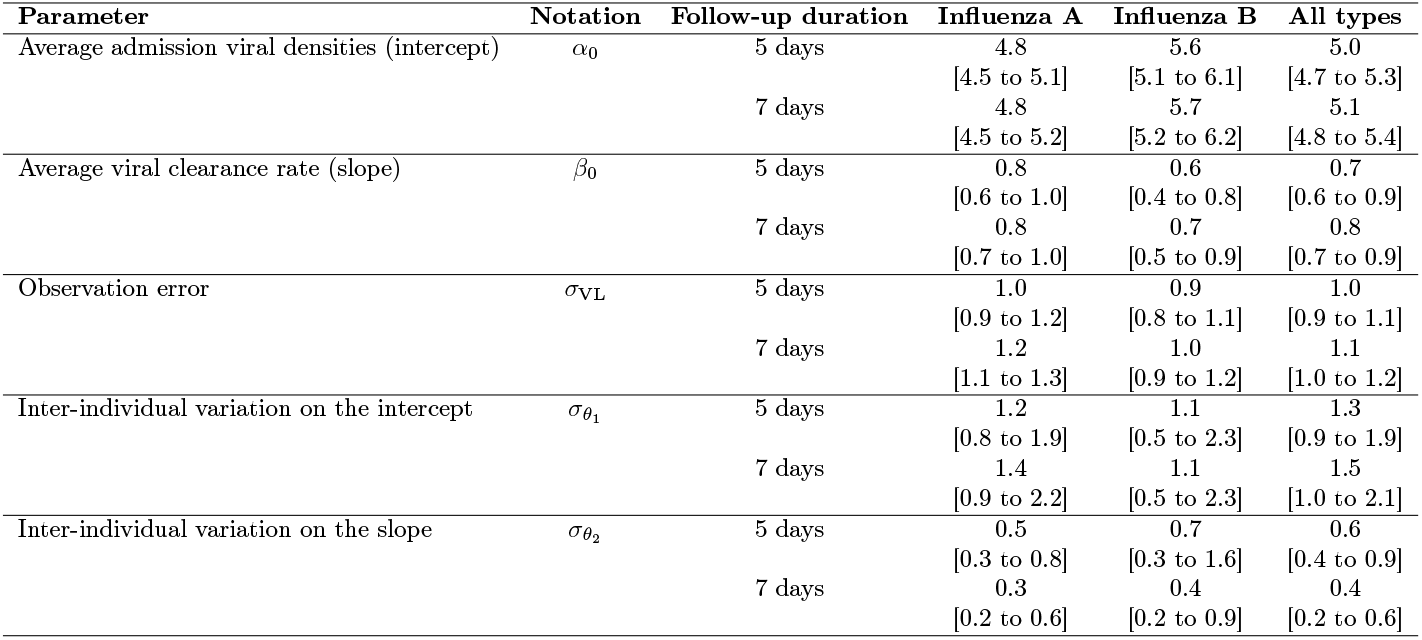
Posterior distributions.

**Table S3:** Sample size per treatment arm required for 90% power, estimated from logistic regression fits.

| Scenario | Effect size |  |  |  |  |
| --- | --- | --- | --- | --- | --- |
|  | 20% | 40% | 60% | 80% | 100% |
| Baseline | 475 | 148 | 80 | 43 | 31 |
| 0.5x intercept variation | 563 | 124 | 52 | 38 | 27 |
| 2.0x intercept variation | 567 | 193 | 66 | 40 | 30 |
| 0.5x slope variation | 149 | 44 | 21 | 14 | 12 |
| 2.0x slope variation | 1981 | 593 | 232 | 148 | 122 |
| 0.5x observation error | 416 | 101 | 60 | 41 | 24 |
| 2.0x observation error | 839 | 162 | 79 | 53 | 35 |
| Sampling every other day (n=3) | 729 | 144 | 80 | 47 | 34 |
| Sampling first and last day (n=2) | 902 | 172 | 111 | 66 | 47 |
| Taking 1 swab/day | 771 | 163 | 81 | 52 | 38 |
| Taking 4 swabs/day | 451 | 127 | 64 | 36 | 26 |

**Table S4:** Sample size per treatment arm required for 80% power, estimated from logistic regression fits.

| Scenario | Effect size |  |  |  |  |
| --- | --- | --- | --- | --- | --- |
|  | 20% | 40% | 60% | 80% | 100% |
| Baseline | 299 | 94 | 52 | 30 | 23 |
| 0.5x intercept variation | 321 | 82 | 37 | 27 | 20 |
| 2.0x intercept variation | 334 | 115 | 44 | 29 | 22 |
| 0.5x slope variation | 93 | 30 | 16 | < 10 | < 10 |
| 2.0x slope variation | 1186 | 364 | 154 | 100 | 77 |
| 0.5x observation error | 250 | 69 | 41 | 28 | 18 |
| 2.0x observation error | 449 | 103 | 51 | 38 | 24 |
| Sampling every other day (n=3) | 400 | 98 | 56 | 34 | 25 |
| Sampling first and last day (n=2) | 444 | 113 | 74 | 46 | 33 |
| Taking 1 swab/day | 431 | 103 | 55 | 37 | 26 |
| Taking 4 swabs/day | 275 | 81 | 45 | 26 | 20 |

**Figure S1:**
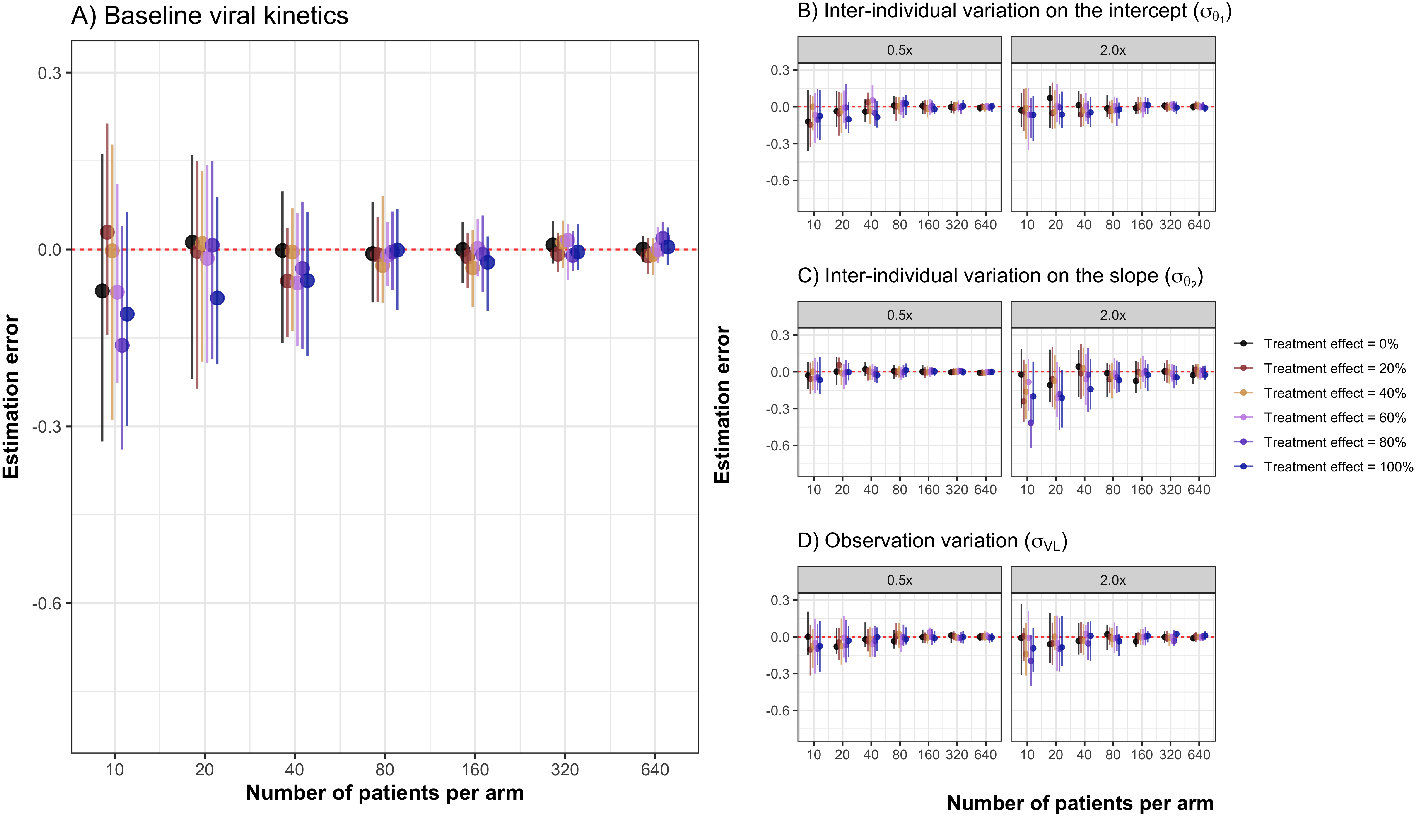
The relationship between the number of patients recruited per arm and the estimation error. Points and error bars represent the median and interquartile range of estimation errors across 50 simulations . (A) Baseline viral kinetics; (B–D) scenarios with half (0.5x) or double (2.0x) the following viral kinetic parameters compared with the baseline (1.0x): (B) observation error (*σ*_*V L*_); (C) inter-individual variation in the intercept 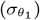; (D) inter-individual variation in the slope 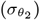.

**Figure S2:**
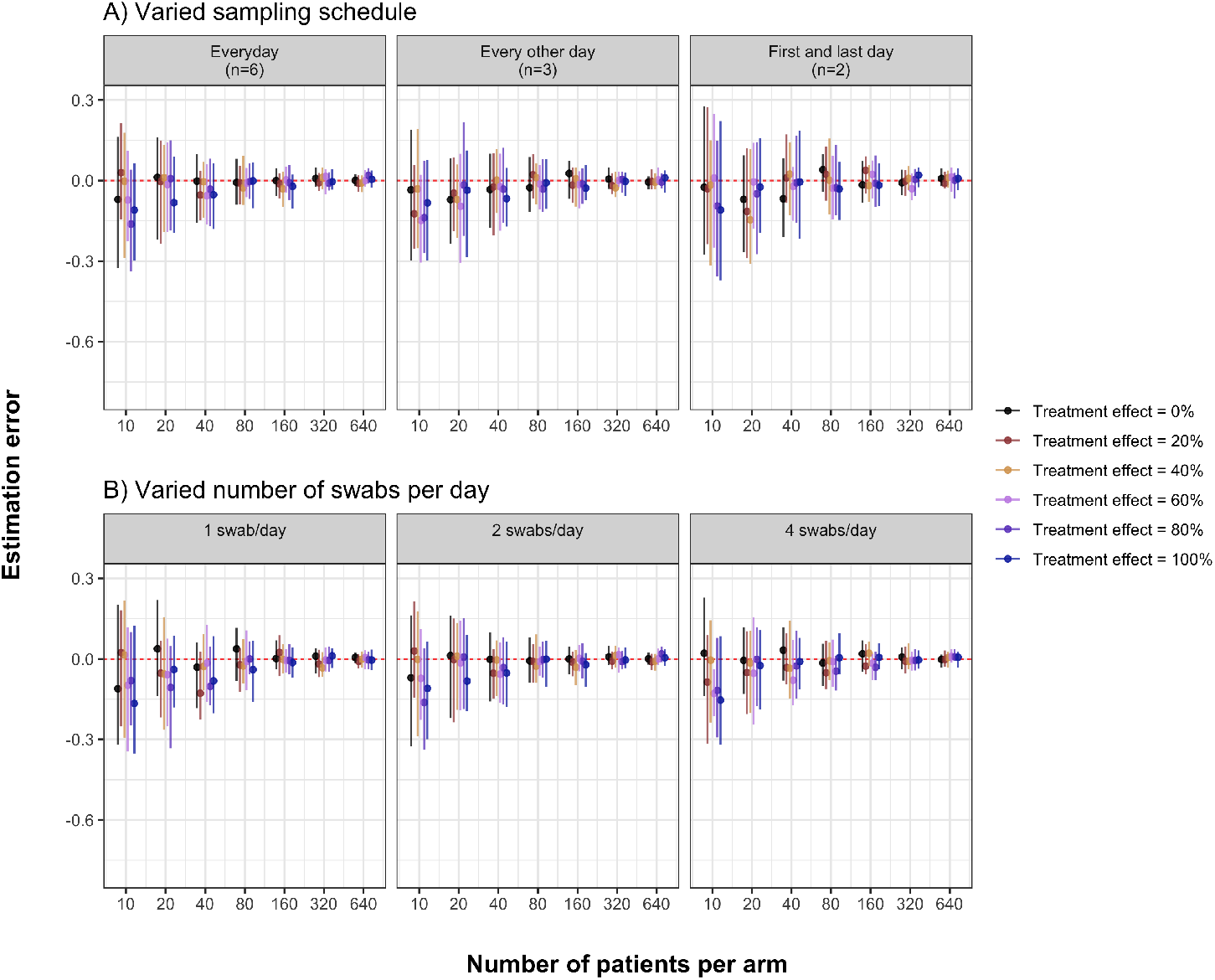
The relationship between the number of patients recruited per arm and the estimation error, using alternative sampling strategies: (A) varied sampling schedule; (B) varied number of swabs per day. Points and error bars represent the median and interquartile range of estimation errors across 50 simulations.

**Figure S3:**
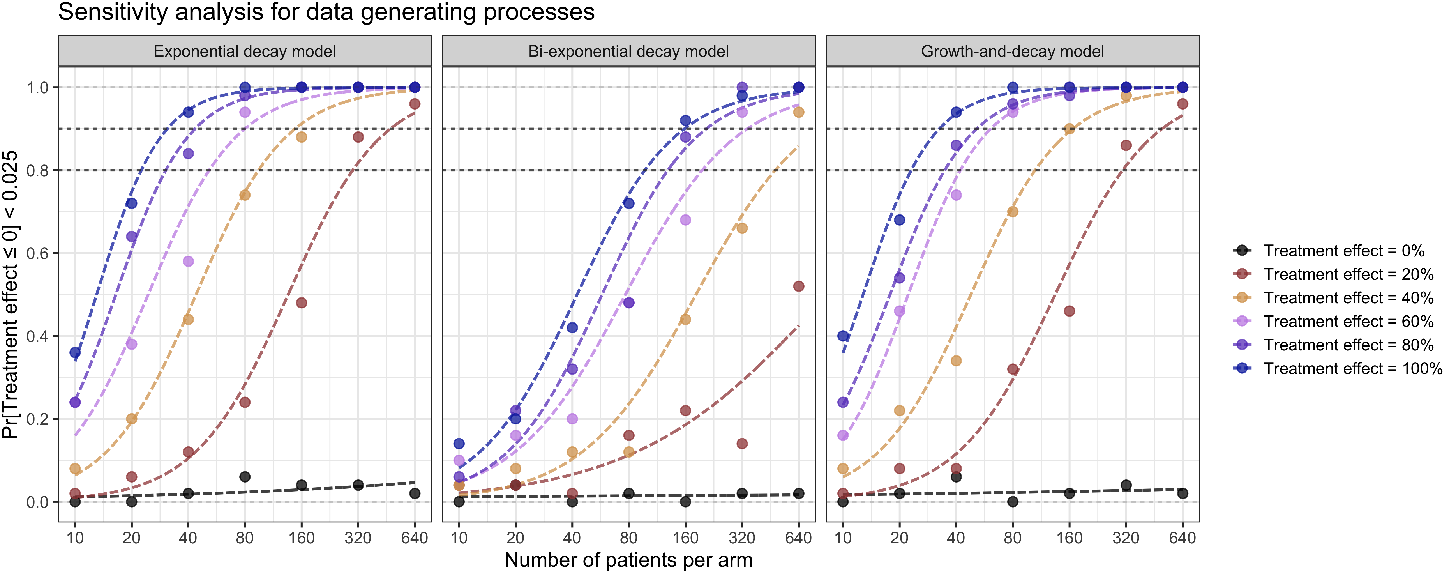
Impact of using different underlying viral kinetic models on the statistical power (or type I error for an effect size of 0%) in simulated clinical trials testing the effectiveness of antiviral treatments for influenza, with colours indicating different treatment effect sizes. Logistic regression fits (dashed lines) are shown to estimate sample sizes required for 80% and 90% power.

