## appendix for "Characterising viral clearance kinetics in acute influenza"

[f] Infectious Diseases Data Observatory, Oxford, United Kingdom.

#### Ethics approval

The trial was approved by the University of Oxford Tropical Research Ethics Committee (OxTREC Ref 6-23) and the Faculty of Tropical Medicine Ethics Committee, Mahidol Univeristy, Thailand (FTEMC Ref TMEC 22-082).

#### Participant inclusion and exclusion criteria

##### Inclusion:

1. Patient understands the procedures and requirements and is willing and able to give informed consent for full participation in the study.
2. Adults, male or female, aged 18 to 60 years at time of consent.
3. Early symptomatic Influenza (A or B); at least one reported symptom of influenza (including fever, history of fever, myalgias, headache, cough, fatigue, nasal congestion, rhinorrhoea and sore throat) within 4 days (96 hours).
4. Influenza positive by rapid antigen test OR a positive RT-PCR test for influenza viruses within the last 24hrs with a Ct value of <30.
5. Able to walk unaided and unimpeded in activities of daily living (ADLs).
6. Agrees and is able to adhere to all study procedures, including availability and contact information for follow-up visits.

##### Exclusion:

1. Taking any concomitant medications or drugs which could interact with the study medications or have antiviral activity
2. Presence of any chronic illness/condition requiring long term treatment or other significant comorbidity
3. BMI ≥35 kg/m$^2$
4. Clinically relevant laboratory abnormalities discovered at screening:
   - Haemoglobin <10 g/dL
   - Platelet count <100,000/µL
   - Alanine transaminase > 1.5 x upper limit of normal
   - Total bilirubin > 1.5 x upper limit of normal
   - Estimated Glomerular Filtration Rate (eGFR) <70 mL/min/1.72 m$^2$
5. For female participants: pregnancy, or actively trying to become pregnant or current breastfeeding

6. Contraindication to taking, or known hypersensitivity reaction to any of the proposed therapeutics
7. Currently participating in another interventional influenza or COVID-19 therapeutic trial
8. Clinical evidence of pneumonia- e.g. shortness of breath, hypoxaemia, crepitations (imaging not required)
9. Known to be currently co-infected with SARS-CoV-2 (i.e. confirmed with positive rapid diagnostic test or qPCR)
10. Received live attenuated influenza virus vaccine within 3 weeks prior to study entry

#### Swabbing procedures

Two oropharyngeal swabs were taken from each tonsil (COPAN Diagnostics, Murrieta, CA, USA). Swabs were rotated 360° against each tonsil four times and placed in viral transport medium (Thermo Fisher, Waltham, MA, USA). Samples were transported at 4-8 °C, aliquoted and frozen at -80 °C. Each tonsil was swabbed daily from enrolment (day 0) to day 7, with an additional swab at day 14. Each swab was processed individually and is treated as a discrete data point in subsequent models.

#### Quantitative PCR procedures

The TaqPath™ one-step RT-qPCR assay and custom complex assay 1TFS-2QAP-CCU002NR (Applied Biosystems, Thermo Fisher Scientific, Waltham, MA, USA) quantitated viral loads (RNA copies per mL). This multiplexed real-time PCR method detects influenza A, B, and human RNase P genes in a single reaction. To quantify viral loads, the ATCC® VR-95DQ A/Puerto Rico/8/1934 (H1N1) strain for influenza A and the ATCC® VR-1804DQ™ B/Florida/4/2006 strain for influenza B were used.

#### Prior distributions

All models are fit using weakly informative priors on all parameters as follows:

- Degree of freedom for Student's t-distribution:
    - $\nu \sim \text{Exponential}(1)$
- Observation error:
    - $\sigma_{VL} \in [0, \infty) \sim \text{Normal}(\text{mean} = 1, \text{sd} = 1)$
- Intercepts (viral densities):
    - $\alpha_0 \sim \text{Normal}(\text{mean} = 5, \text{sd} = 2)$
    - $\alpha_1 \sim \text{Normal}(\text{mean} = 5, \text{sd} = 2)$
    - $\alpha_2 \sim \text{Normal}(\text{mean} = 3, \text{sd} = 2)$
- Slopes (clearance/growth rate):
    - $\beta_0 \sim \text{Normal}(\text{mean} = 0.5, \text{sd} = 1)$
    - $\beta_1 \sim \text{Normal}(\text{mean} = 1, \text{sd} = 1)$
    - $\beta_2 \sim \text{Normal}(\text{mean} = 0.5, \text{sd} = 1)$
- Time at peak viral densities:
    - $\tau_{\text{max}} \sim \text{Normal}(\text{mean} = -3, \text{sd} = 3)$
- Individual random effects $\theta_i$ have a multivariate normal distribution with Cholesky parameterization as a prior, with location parameter $\mu = 0$ and standard deviation $\Sigma \sim \text{Exponential}(1)$ and correlation matrix for individual random effects $\Omega \sim \text{Cholesky}(2)$ as hyperparameters.

### Individual patient viral dynamics plots

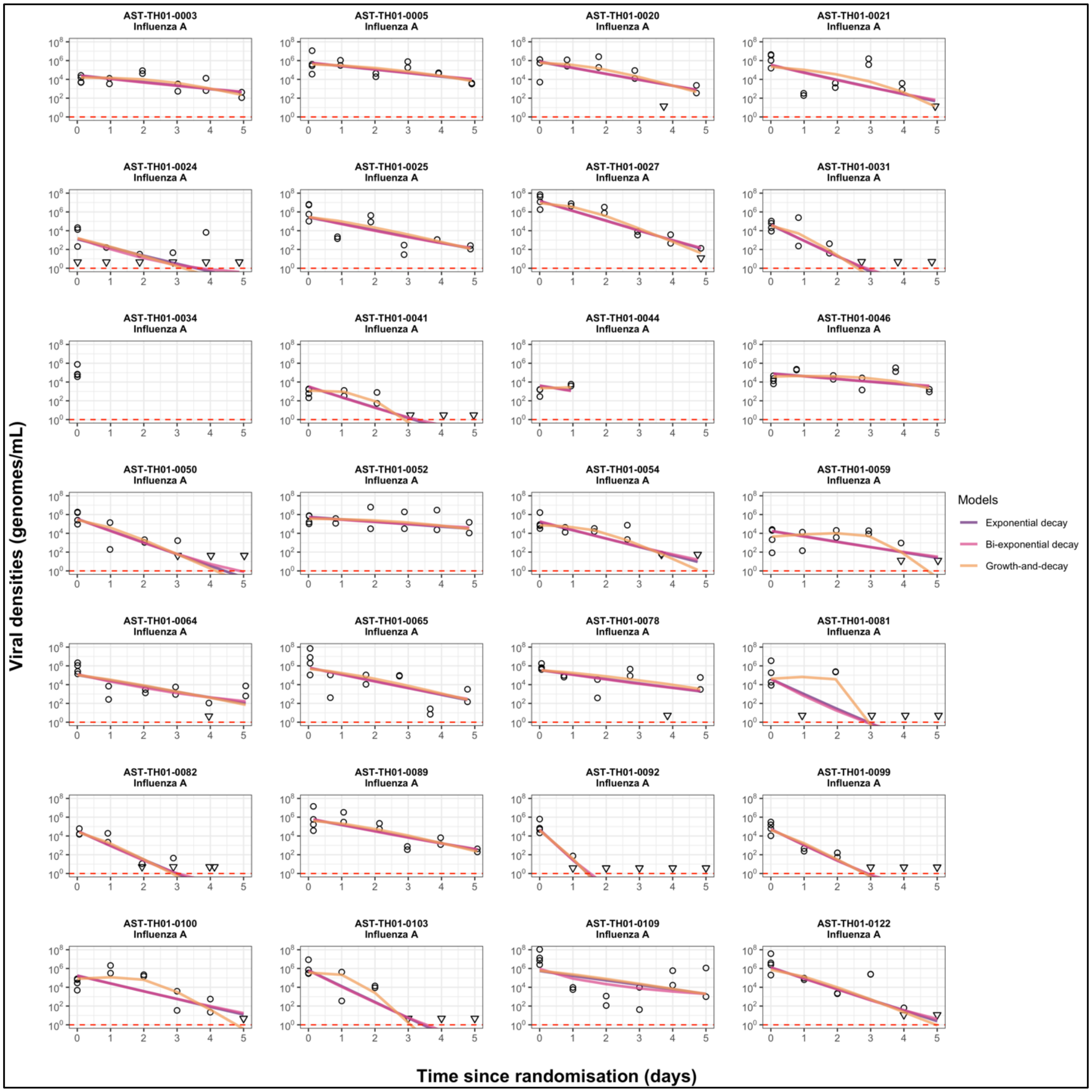

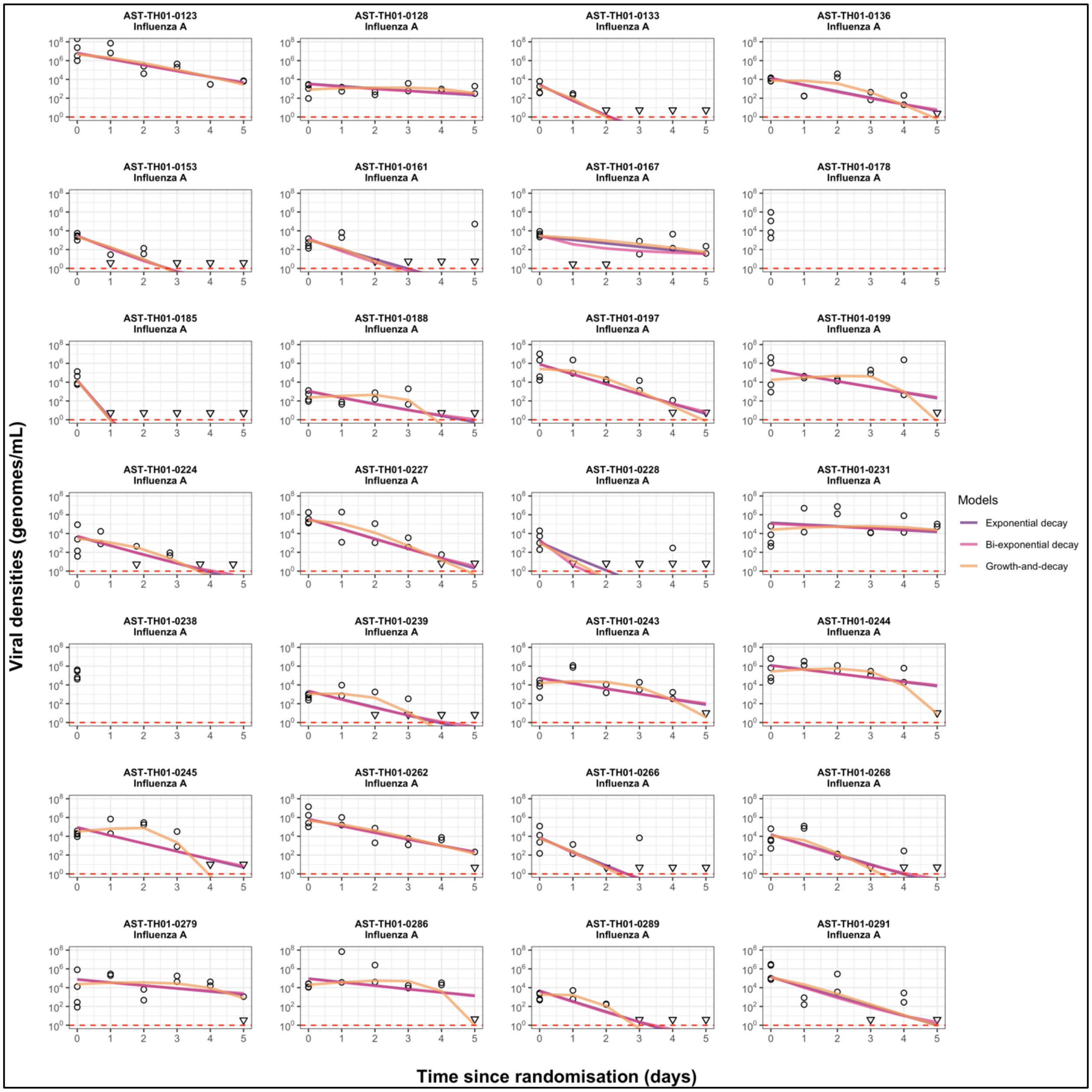
AST-TH01-0123 Influenza A
AST-TH01-0128 Influenza A
AST-TH01-0133 Influenza A
AST-TH01-0136 Influenza A
AST-TH01-0153 Influenza A
AST-TH01-0161 Influenza A
AST-TH01-0167 Influenza A
AST-TH01-0178 Influenza A
AST-TH01-0185 Influenza A
AST-TH01-0188 Influenza A
AST-TH01-0197 Influenza A
AST-TH01-0199 Influenza A
AST-TH01-0224 Influenza A
AST-TH01-0227 Influenza A
AST-TH01-0228 Influenza A
AST-TH01-0231 Influenza A
AST-TH01-0238 Influenza A
AST-TH01-0239 Influenza A
AST-TH01-0243 Influenza A
AST-TH01-0244 Influenza A
AST-TH01-0245 Influenza A
AST-TH01-0262 Influenza A
AST-TH01-0266 Influenza A
AST-TH01-0268 Influenza A
AST-TH01-0279 Influenza A
AST-TH01-0286 Influenza A
AST-TH01-0289 Influenza A
AST-TH01-0291 Influenza A
Viral densities (genomes/mL)
Time since randomisation (days)
Models
Exponential decay
Bi-exponential decay
Growth-and-decay

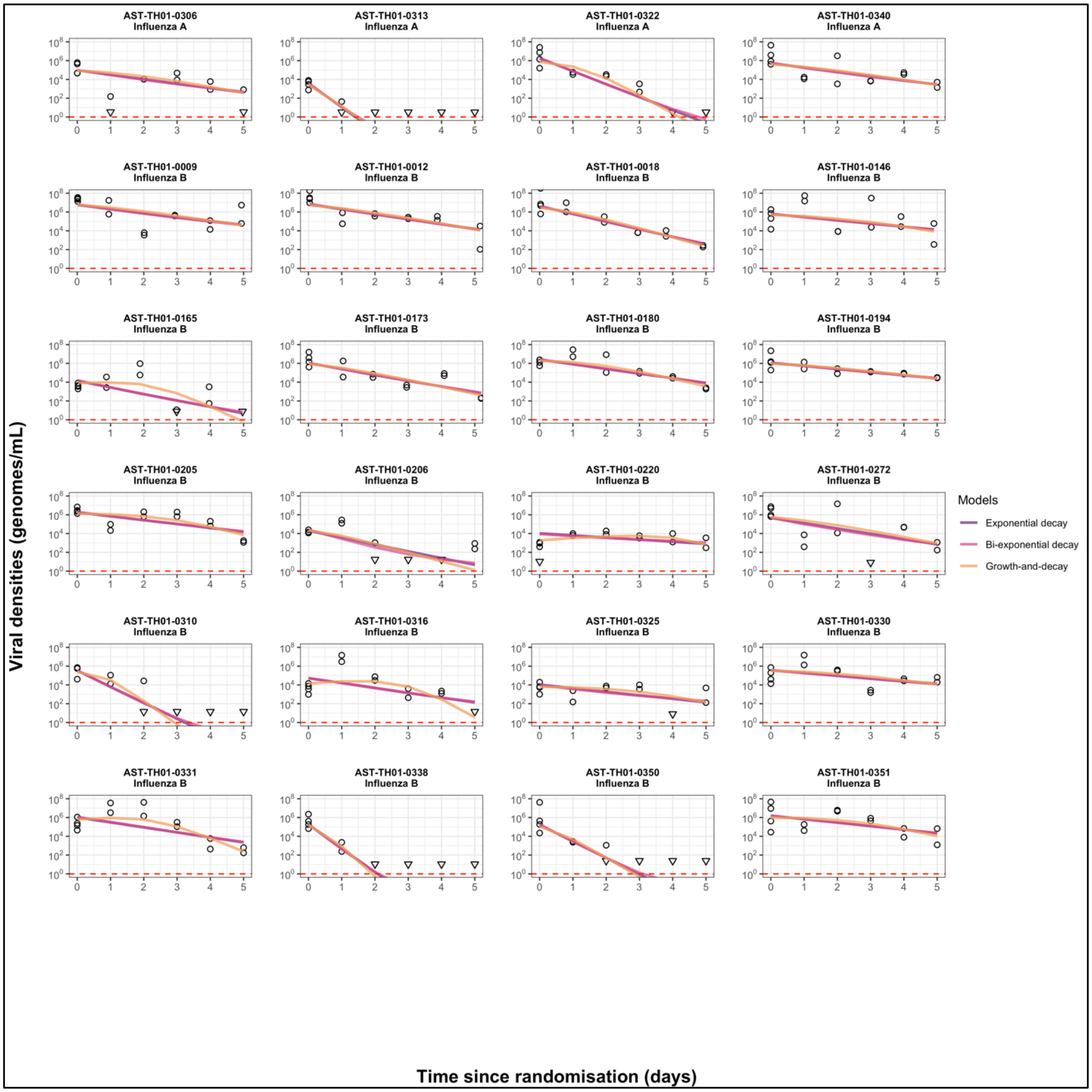
AST-TH01-0306
Influenza A
AST-TH01-0313
Influenza A
AST-TH01-0322
Influenza A
AST-TH01-0340
Influenza A
AST-TH01-0009
Influenza B
AST-TH01-0012
Influenza B
AST-TH01-0018
Influenza B
AST-TH01-0146
Influenza B
AST-TH01-0165
Influenza B
AST-TH01-0173
Influenza B
AST-TH01-0180
Influenza B
AST-TH01-0194
Influenza B
AST-TH01-0205
Influenza B
AST-TH01-0206
Influenza B
AST-TH01-0220
Influenza B
AST-TH01-0272
Influenza B
AST-TH01-0310
Influenza B
AST-TH01-0316
Influenza B
AST-TH01-0325
Influenza B
AST-TH01-0330
Influenza B
AST-TH01-0331
Influenza B
AST-TH01-0338
Influenza B
AST-TH01-0350
Influenza B
AST-TH01-0351
Influenza B
Viral densities (genomes/mL)
Time since randomisation (days)
Models
Exponential decay
Bi-exponential decay
Growth-and-decay
